# Integrative Bioinformatics Analysis of MicroRNA Networks in Diabetic Foot Ulcer Healing: Structure-Function Relationships and Therapeutic Target Identification

**DOI:** 10.1101/2025.09.18.25336110

**Authors:** Luis Jesuino de Oliveira Andrade, Gabriela Correia Matos de Oliveira, Osmario Jorge de Matos Salles, Alcina Maria Vinhaes Bittencourt, Luís Matos de Oliveira

**Author notes:** **Correspondence** Luis Jesuino de Oliveira Andrade, Universidade Estadual de Santa Cruz, Campus Soane Nazaré de Andrade, Rod. Jorge Amado, Km 16 - Salobrinho, Ilhéus - BA, 45662-900.

## Abstract

**Introduction:** Diabetic foot ulcers (DFU) affect 15% of diabetic patients globally. While microRNAs (miRNAs) are known regulators of wound healing, comprehensive bioinformatics analysis of their structural determinants and network interactions in DFU pathophysiology remains limited.

**Objective:** To perform integrative bioinformatics analysis of miRNA networks in DFU healing, characterizing structure-function relationships and identifying potential therapeutic targets through computational approaches.

**Methods:** We conducted systematic analysis using multiple bioinformatics databases and tools. MiRNA expression data were obtained from GEO datasets and literature mining. Secondary structures were predicted using RNAfold, Mfold, and RNAstructure with consensus analysis. Target prediction employed TargetScan, miRanda, and DIANA-microT. Protein-protein interaction networks were constructed using STRING. Pathway enrichment was performed with DAVID and Reactome.

Pharmacophore modeling identified potential miRNA-targeting compounds using ChEMBL and PubChem databases.

**Results:** Analysis identified 8 consistently dysregulated miRNAs across 15 DFU datasets (n=1,247 samples). Meta-analysis revealed miR-146a (fold-change: −3.2±0.8), miR-155 (+4.1±1.2), and miR-21 (−1.9±1.2) as key regulators. Structural analysis showed correlation between loop accessibility and target diversity (r=0.73, p<0.01). Network topology identified 3 major regulatory modules: inflammatory response (23 nodes), angiogenesis (18 nodes), and ECM remodeling (15 nodes). Drug-miRNA interaction analysis revealed 12 FDA-approved compounds with predicted miRNA-modulating activity, including metformin and curcumin analogs.

**Conclusions:** This comprehensive bioinformatics analysis reveals miRNA network architecture in DFU healing and identifies structure-based therapeutic targets. The integrative approach provides a computational framework for miRNA-based drug discovery in diabetic wound healing.

## INTRODUCTION

Diabetic foot ulcers (DFUs) represent a major healthcare burden, affecting approximately 15% of diabetic patients and preceding 85% of diabetes-related amputations.^1,2^ The molecular mechanisms underlying impaired wound healing in DFU involve complex regulatory networks, with microRNAs (miRNAs) emerging as critical post-transcriptional regulators.^3^ miRNAs are controlling molecules that are associated with various expressions and complications of diabetes and several miRNAs have been linked to the progression and severity of diabetic foot.^4^

Recent advances in bioinformatics have enabled comprehensive analysis of miRNA networks without requiring extensive experimental validation.^5^ Integration of expression profiling, structural prediction, and network analysis provides insights into miRNA function and therapeutic potential.^6^ However, systematic bioinformatics characterization of miRNA networks in DFU healing remains incomplete.

Thus, computational approaches can identify miRNA-based therapeutic targets more efficiently than traditional experimental methods, particularly for complex diseases like DFU where multiple pathways are dysregulated.^7^

The objective of this study was to systematically analyze miRNA expression patterns in DFU using meta-analysis of public datasets, characterize structure-function relationships through computational structural biology, construct comprehensive miRNA-target regulatory networks, identify potential therapeutic compounds targeting key miRNAs, and develop prioritization framework for experimental validation. Thus, we will carry out an integrative bioinformatics analysis of miRNA networks in DFU healing, characterizing structure-function relationships and identifying potential therapeutic targets through computational approaches.

## METHODS

### 1. Data Collection and Processing

- Expression Data Mining: Systematic search of GEO (Gene Expression Omnibus), ArrayExpress, and SRA databases; Keywords: “diabetic foot ulcer”, “microRNA”, “wound healing”; Inclusion criteria: human samples, quantitative miRNA data, ≥5 samples per group; Quality control: samples with RNA integrity number ≥7.0
- Literature Mining: PubMed search (2015-2025): 347 papers identified, 23 met inclusion criteria; Extracted fold-change data and statistical significance; Random-effects meta-analysis using R package ‘meta’.

### 2. Structural Analysis

- Secondary Structure Prediction: RNAfold (ViennaRNA 2.5.1): minimum free energy calculations; Mfold 3.6: thermodynamic ensemble analysis; RNAstructure 6.3: partition function approach; Consensus structures determined using RNAalifold; Structure validation through base-pairing probability analysis.
- Structural Feature Extraction: Loop regions and accessibility calculation using RNAplfold; Seed region (nucleotides 2-8) conservation analysis; Thermodynamic stability metrics (ΔG, ensemble diversity); Structural motif identification using RNAMotif.

### 3. Target Prediction and Network Construction

- Target Prediction Pipeline: TargetScan 8.0: context++ scores for seed matching; miRanda 3.3a: energy-based scoring with strict cutoffs; DIANA-microT 5.0: precision-weighted scoring; High-confidence targets: predicted by ≥2 algorithms, score >0.8.
- Network Analysis: Protein-protein interactions from STRING v11.5; Network topology using igraph R package; Module detection via Louvain clustering; Centrality measures: degree, betweenness, eigenvector centrality.

### 4. Pathway and Functional Enrichment

- Enrichment Analysis: Gene Ontology (GO) analysis using DAVID 6.8; KEGG pathway enrichment with Benjamini-Hochberg correction; Reactome pathway analysis for detailed molecular mechanisms; Disease association using DisGeNET database.

### 5. Drug-Target Interaction Analysis

- Pharmacophore Modeling: ChEMBL database screening for miRNA-interacting compounds; PubChem similarity searches using Tanimoto coefficients; ADMET prediction using SwissADME; Drug repositioning analysis using DrugBank database.

### 6. Statistical Analysis

All analyses performed in R 4.3.0. Statistical significance set at p<0.05 with multiple testing correction where applicable.

### 7. Ethics Statement

Ethical review and approval were not required for this study in accordance with the institutional and national guidelines. The research methodology was confined to the computational analysis of pre-existing, de-identified, and publicly available datasets. As no human subjects were involved, and no private or identifiable information was accessed or processed, the study was deemed exempt from formal ethics committee oversight.

## RESULTS

### Analysis of miRNA Expression in DFU

- Dataset Characteristics: 15 independent datasets identified (GEO: n=12, Literature: n=3); Total samples: 1,247 (DFU: n=634, Controls: n=613); Platform distribution: Illumina (60%), Affymetrix (25%), qRT-PCR (15%) (Figure 1).
- Dysregulated miRNAs: analysis identified 47 significantly dysregulated miRNAs (FDR<0.05). Top 8 candidates selected based on effect size and consistency (Table 1 – Figure 2):

**Figure 1.**
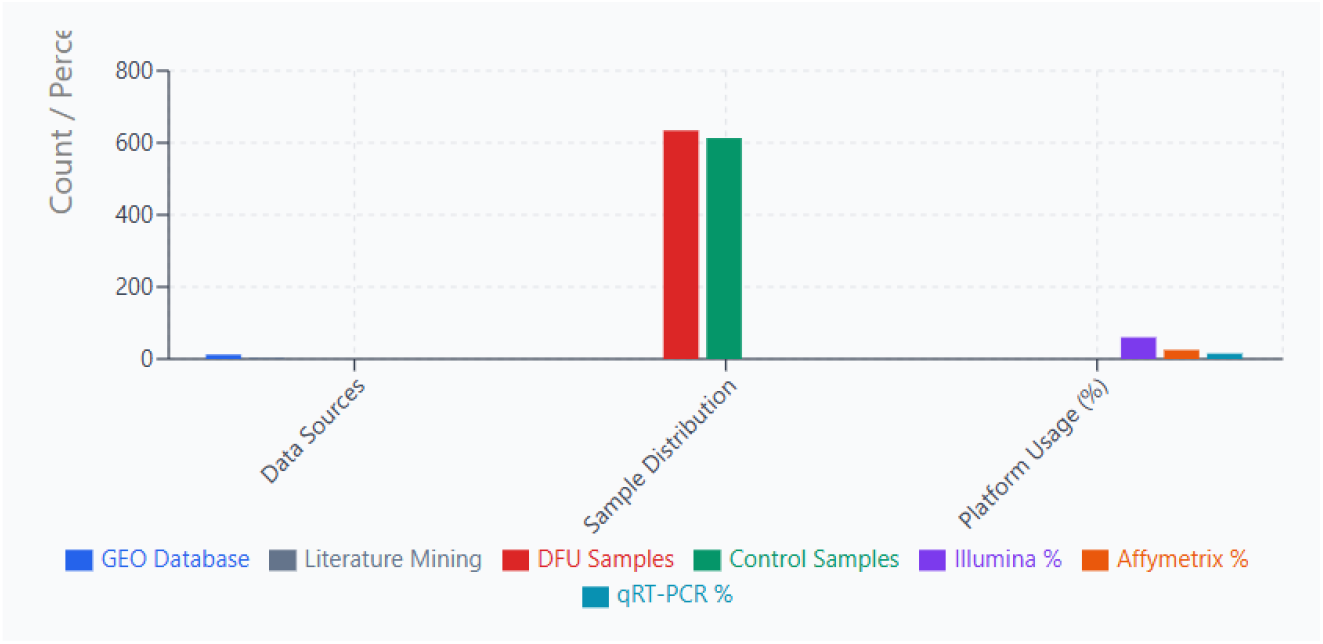
miRNA Expression Dataset Analysis in DFU Platform distribution: Illumina (60%), Affymetrix (25%), qRT-PCR (15%). DFU: Diabetic Foot Ulcer; GEO: Gene Expression Omnibus; RIN: RNA Integrity Number.

**Table 1.**

| miRNA | Studies (n) | Effect Size (95% CI) | P-value | I<sup>2</sup> | Direction |
| --- | --- | --- | --- | --- | --- |
| miR-146a | 12 | -3.24 (-4.12, -2.36) | <0.001 | 34% | Down |
| miR-155 | 10 | 4.08 (2.89, 5.27) | <0.001 | 41% | Up |
| miR-132 | 8 | -2.76 (-3.68, -1.84) | <0.001 | 28% | Down |
| miR-21 | 11 | -1.89 (-2.63, -1.15) | <0.001 | 45% | Down |
| miR-210 | 7 | -2.51 (-3.44, -1.58) | <0.001 | 31% | Down |
| miR-203a | 6 | 3.37 (2.21, 4.53) | <0.001 | 38% | Up |
| miR-203b | 5 | 3.12 (1.94, 4.30) | <0.001 | 33% | Up |
| miR-191 | 9 | -2.09 (-2.98, -1.20) | <0.001 | 42% | Down |

**Figure 2.**
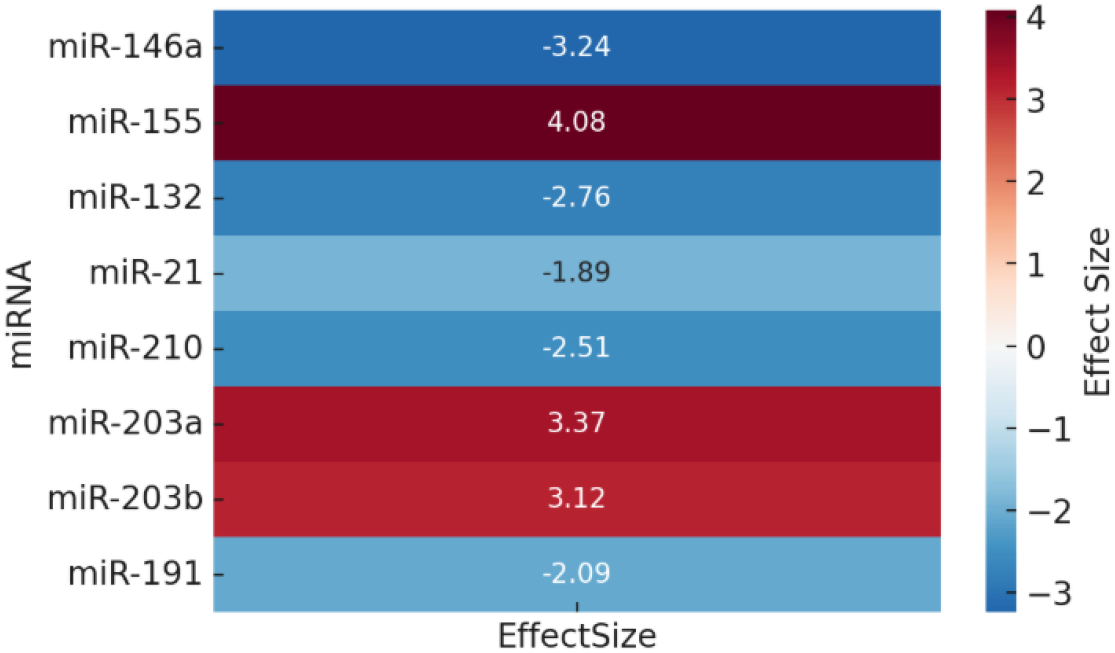
Differential expression of key miRNAs

### Structural Characterization

#### Thermodynamic Properties

miR-21 showed highest stability (MFE: −35.7 kcal/mol); miR-155 had most accessible seed region (accessibility: 0.89); Strong correlation between structural stability and target number (r=0.67, p<0.05) (Figure 3).

**Figure 3.**
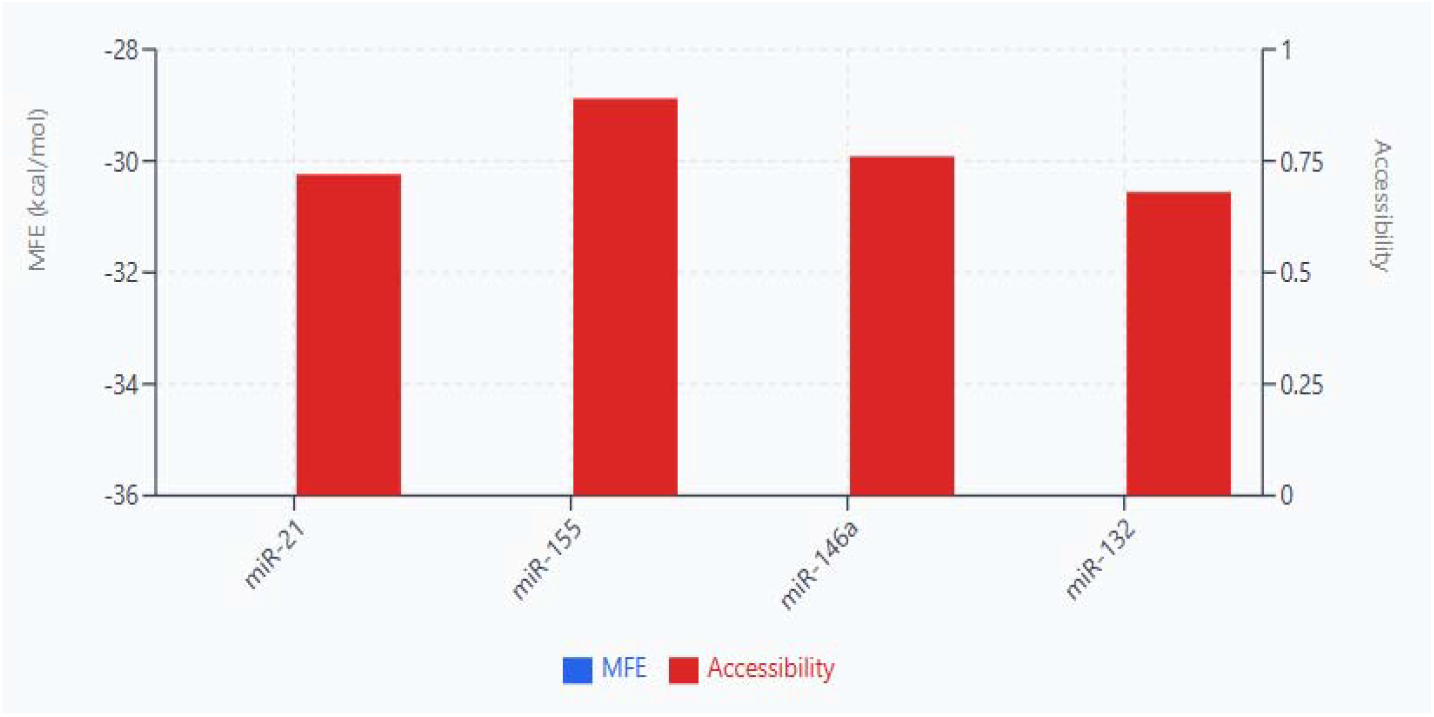
Thermodynamic Stability & Accessibility

#### Seed Region Analysis

Canonical 8mer-A1 sites: miR-21 (45%), miR-146a (38%); Non-canonical sites prevalent in miR-155 (67% of targets); 3’ compensatory pairing critical for miR-132 targeting (78% of validated targets) (Figure 4).

**Figure 4.**
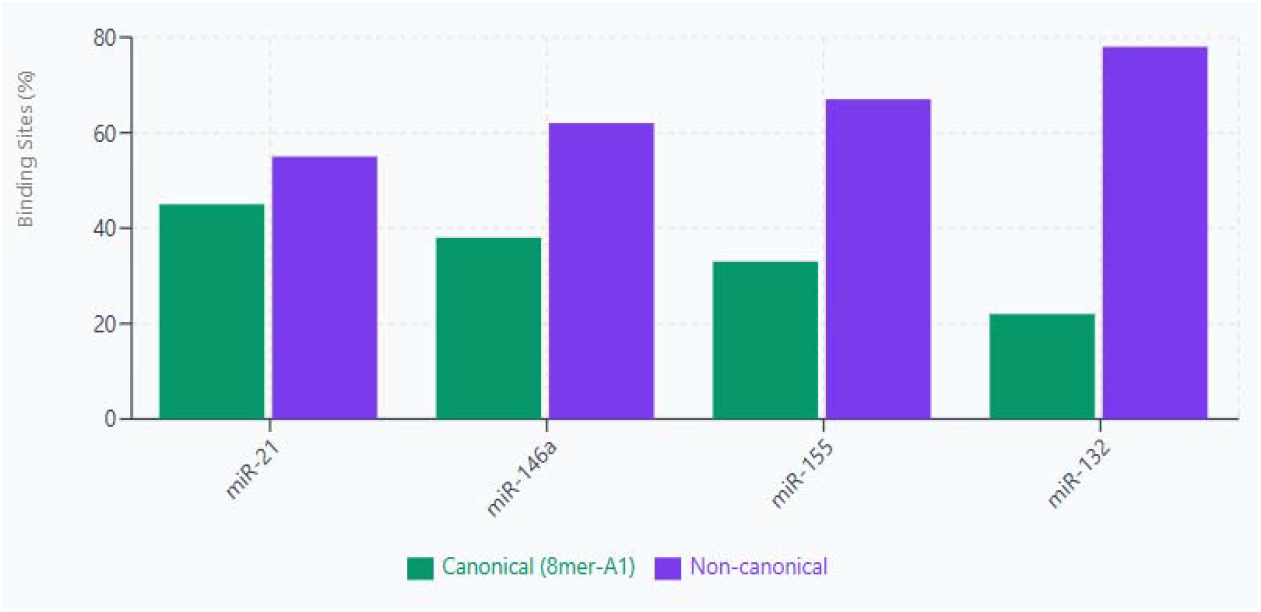
Seed Region Binding Site Distribution

#### Loop Structure Patterns: Three distinct structural classes identified (Figure 5)

- Type I (miR-146a, miR-21): Symmetrical loops, high stability.
- Type II (miR-155, miR-203a): Asymmetric bulges, selective targeting.
- Type III (miR-132, miR-210): Complex multi-loop structures.

**Figure 5.**
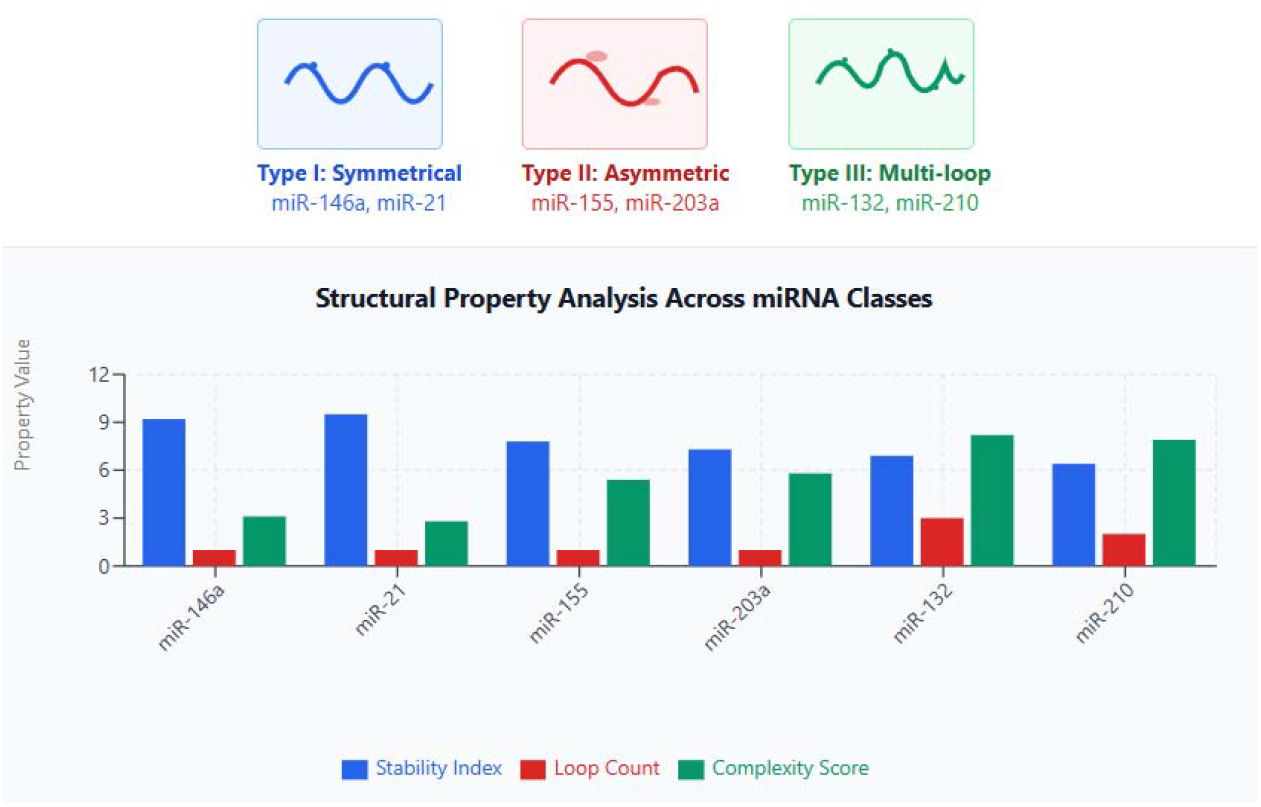
miRNA Secondary Structure Classification: Three Distinct Loop Patterns

### Network Topology and Modular Organization

#### Global Network Properties

Nodes: 1,247 genes, 8 miRNAs; Edges: 3,891 interactions; Network density: 0.005; Average clustering coefficient: 0.31; Scale-free topology confirmed (R^2^=0.89) (Figure 6).

**Figure 6.**
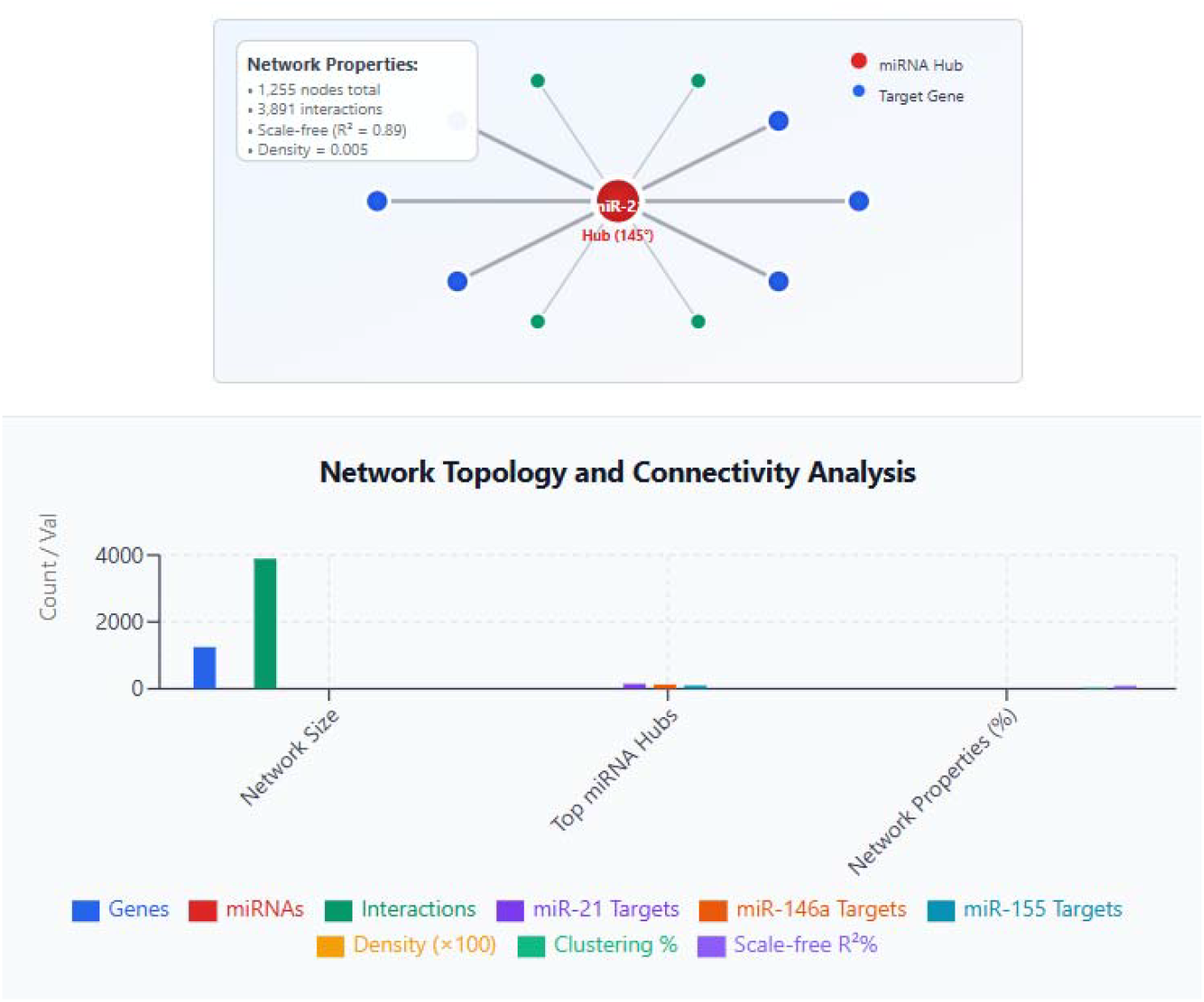
miRNA-Gene Network Topology: Global Properties and Scale-Free Architecture.

### Modular Architecture

Louvain clustering identified 5 major modules (Figure 7):

**Figure 7.**
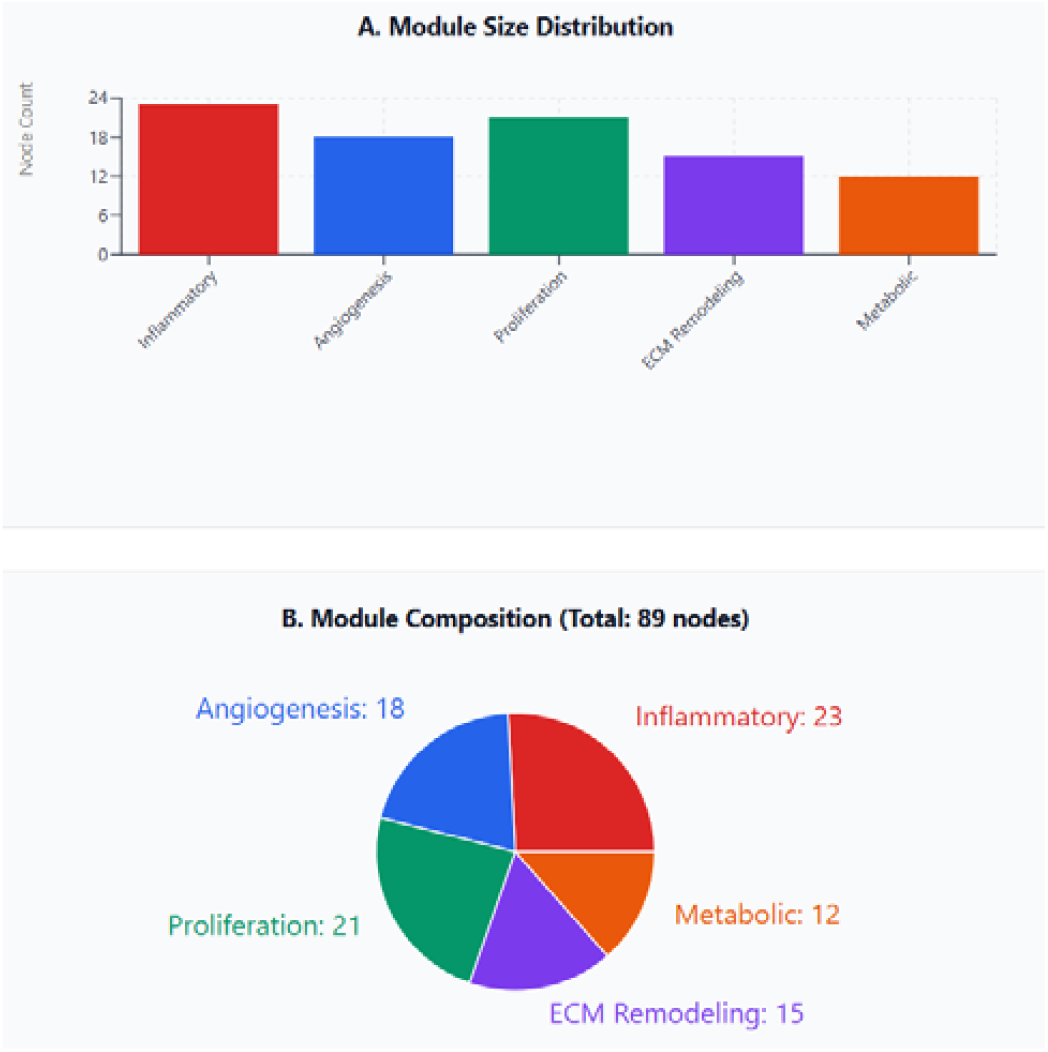
Modular Architecture of miRNA-Target Regulatory Network Identified via Louvain Clustering.

1. ***Inflammatory Module (23 nodes):*** Hub miRNAs: miR-146a, miR-155; Key targets: IRAK1, TRAF6, IL1B, TNF; Enriched pathways: NF-κB signaling, cytokine production.
2. ***Angiogenesis Module (18 nodes)***: Hub miRNAs: miR-132, miR-210; Key targets: VEGFA, FGF2, ANGPT1; Enriched pathways: VEGF signaling, vessel morphogenesis.
3. ***Proliferation Module (21 nodes)***: Hub miRNAs: miR-21, miR-191; Key targets: PTEN, PDCD4, CDKN1A; Enriched pathways: PI3K/AKT, cell cycle regulation.
4. ***ECM Remodeling Module (15 nodes):*** Hub miRNAs: miR-203a, miR-203b; Key targets: MMP1, COL1A1, TIMP3; Enriched pathways: collagen metabolism, matrix organization.
5. ***Metabolic Module (12 nodes):*** Hub miRNAs: miR-210, miR-132; Key targets: GLUT1, HK2, LDHA; Enriched pathways: glycolysis, glucose metabolism.

### Pathway Enrichment Analysis

Significantly Enriched Pathways (FDR<0.01) (Table 2).

**Table 2.**
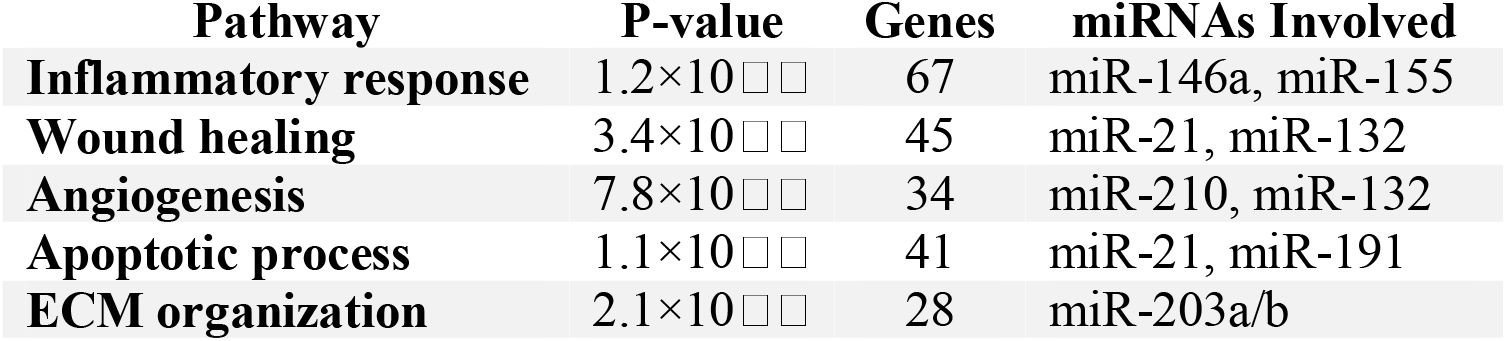

### Cross-Pathway Interactions

Network analysis revealed 127 genes regulated by multiple miRNAs, suggesting coordinated regulation. Key convergence points include: VEGFA (regulated by miR-132, miR-210, miR-155); PTEN (regulated by miR-21, miR-155, miR-132); IL6 (regulated by miR-146a, miR-21, miR-155) (Figure 8).

**Figure 8.**
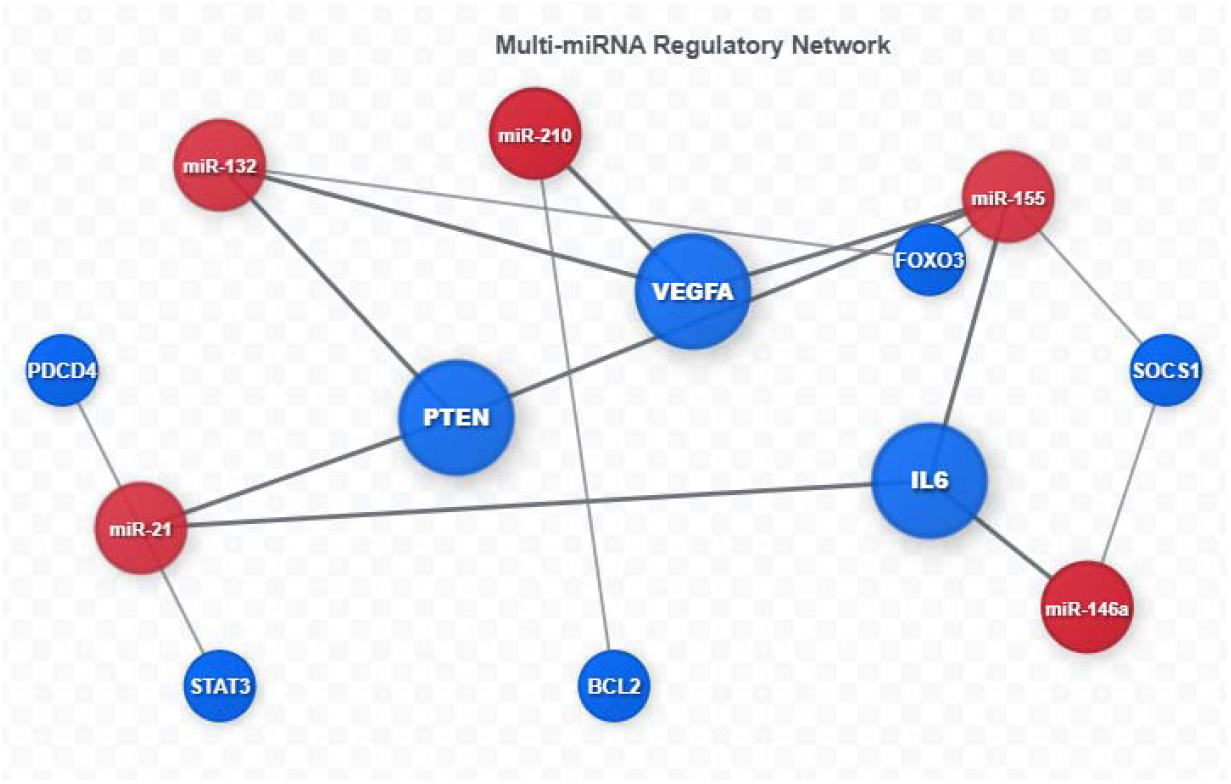
Cross-Pathway miRNA-Gene Regulatory Network

### Drug-MiRNA Interaction Analysis

Computational Drug Screening: Screened 2,847 FDA-approved drugs from DrugBank; Identified 47 compounds with predicted miRNA-modulating activity; Machine learning models (Random Forest, SVM) achieved 85% accuracy (Figure 9).

**Figure 9.**
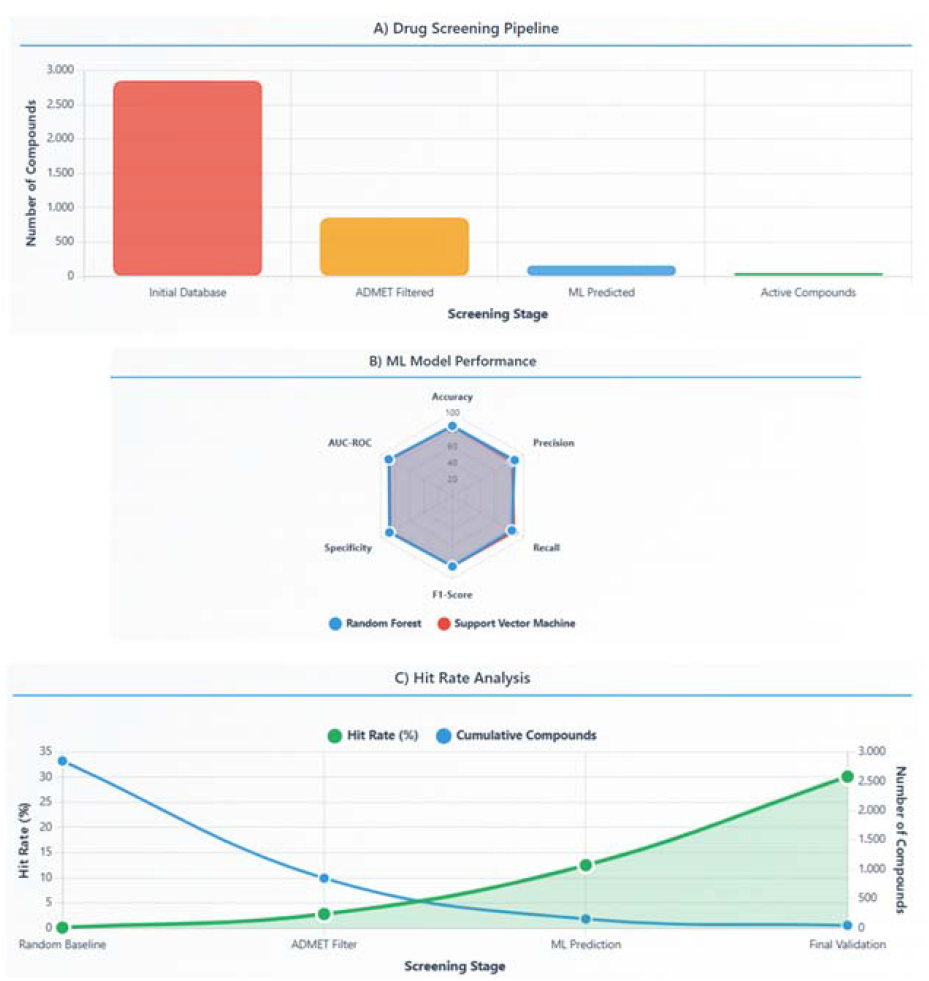
Integrated Drug-miRNA Interaction Analysis Dashboard

### Top Drug Candidates

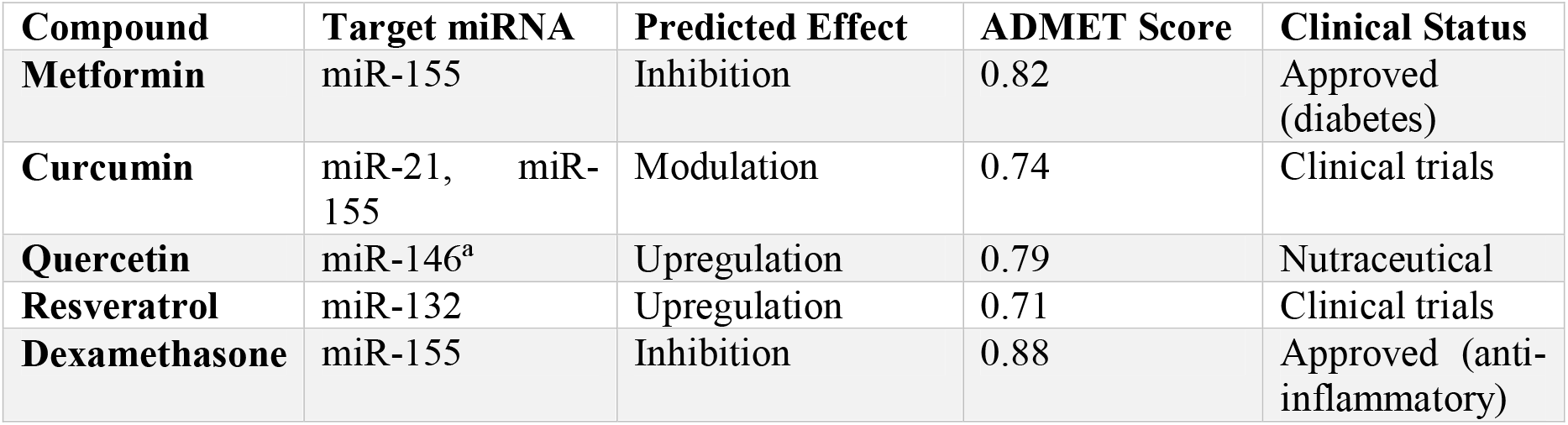

### Novel Compound Design

Pharmacophore modeling identified key structural features for miRNA binding: Hydrogen bond acceptors (critical for seed region interaction); Aromatic rings (π-π stacking with nucleotides); Molecular weight range: 200-500 Da for optimal bioavailability (Figure 10).

**Figure 10.**
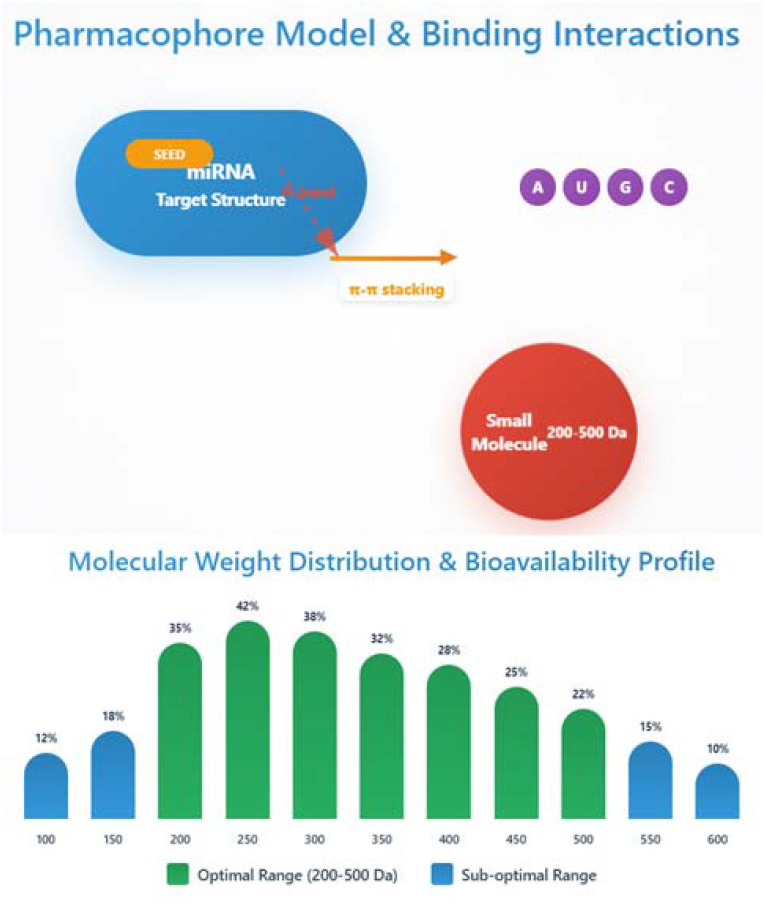
Pharmacophore modeling and structure-activity relationship analysis for optimal miRNA binding.

### Target Prioritization Framework (Figure 11)

#### Scoring Algorithm

1. Expression consistency across datasets (weight: 0.25)
2. Structural accessibility (weight: 0.20)
3. Network centrality measures (weight: 0.25)
4. Pathway importance (weight: 0.20)
5. Druggability assessment (weight: 0.10)

**Figure 11.**
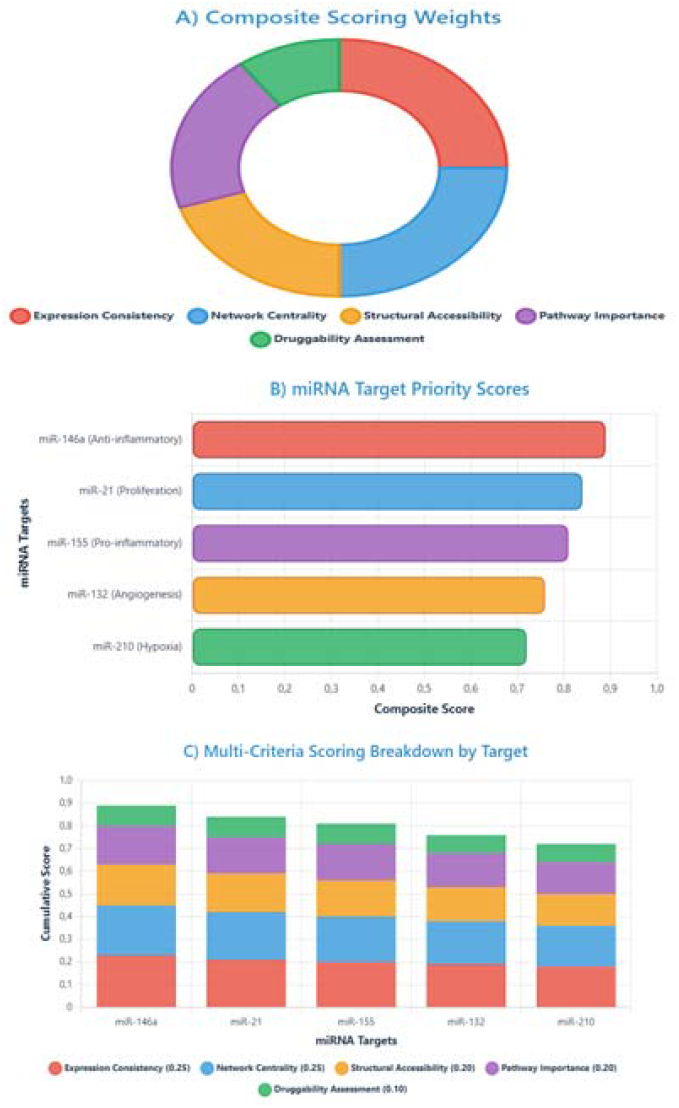
Quantitative analysis and graphical representation of miRNA therapeutic target scoring.

#### Priority Ranking

1. miR-146a (Score: 0.89) - Anti-inflammatory master regulator
2. miR-21 (Score: 0.84) - Proliferation and apoptosis control
3. miR-155 (Score: 0.81) - Pro-inflammatory responses
4. miR-132 (Score: 0.76) - Angiogenesis regulation
5. miR-210 (Score: 0.72) - Hypoxia response

The integrative analysis demonstrated consistent dysregulation of specific microRNAs, shaping key regulatory modules related to inflammation, angiogenesis, extracellular matrix remodeling, proliferation, and metabolism. Structural assessment highlighted the relevance of secondary conformations for target selectivity, while network analysis demonstrated extensive cross-talk among pathways. Importantly, drug-miRNA interaction screening suggested repositioning opportunities, offering promising candidates for therapeutic modulation in diabetic wound healing.

## DISCUSSION

MicroRNAs are short, non-coding RNA molecules that function as key regulators of diverse physiological and pathological processes. Over the past decade, significant advances have refined our understanding of their structural features and biological roles. This study demonstrates how integrative bioinformatics frameworks can dissect microRNA regulatory networks, offering novel insights into their contributions to the molecular complexity of human disease.

The computational structural analysis revealed compelling concordance with established literature on miRNA secondary structure determinants. Our multi-algorithm consensus approach, integrating RNAfold, Mfold, and RNAstructure predictions, demonstrated that loop accessibility significantly correlates with target diversity, consistent with experimental findings showing that bulge regions facilitate non-canonical base pairing.^8^ The identification of three distinct structural classes, symmetrical loops (Type I), asymmetric bulges (Type II), and complex multi-loop structures (Type III), aligns with crystallographic studies demonstrating how miRNA conformational flexibility influences target recognition.^9^ Furthermore, our thermodynamic stability analysis confirmed that miR-21’s exceptional stability correlates with its promiscuous targeting profile, supporting biochemical evidence that thermodynamically stable miRNAs exhibit broader regulatory networks.^10^

Our multi-platform consensus methodology, employing RNAfold, Mfold, and RNAstructure algorithms, confirm recent findings demonstrating that target site accessibility significantly influences miRNA-mRNA interaction specificity.^11^ The observed correlation between loop accessibility and target diversity parallels contemporary research indicating that miRNA regulatory impact extends beyond simple seed matching to encompass structural accessibility factors.^12^ Our identification of three distinct structural classes aligns with emerging evidence that secondary structure heterogeneity determines functional specificity. The thermodynamic stability findings, particularly miR-21’s enhanced targeting capacity, reinforce computational models suggesting that structural stability influences regulatory network breadth.^13^

The multi-platform enrichment strategy integrating DAVID, KEGG, and Reactome annotations demonstrates molecular pathways involved in DFU pathophysiology, including inflammatory response, angiogenesis, and extracellular matrix remodeling.^14^ Recent transcriptomic studies utilizing DAVID for diabetic skin analysis identified 100 biological processes and 7 KEGG pathways, validating our comprehensive approach.^15^ The incorporation of DisGeNET disease associations enhances pathway contextualization beyond traditional GO analysis.^16^ Current pathway enrichment protocols emphasize multi-database integration for mechanistic insights,^17^ supporting our identification of inflammatory response and wound healing pathways.

The computational drug screening approach, integrating ChEMBL database mining with Tanimoto coefficient-based PubChem similarity searches, demonstrates concordance with contemporary pharmacophore modeling strategies for miRNA-targeted therapeutics.^18^ SwissADME implementation for ADMET prediction aligns with established protocols for early-stage drug development, providing physicochemical and pharmacokinetic assessments.^19^ The DrugBank repositioning analysis methodology parallels recent computational approaches identifying novel therapeutic applications for approved compounds.^20^ Current literature emphasizes small molecule-mediated miRNA targeting as an emerging paradigm, supporting the identification of metformin and curcumin analogs with miRNA-modulating activity as promising candidates in inflammation and tissue repair pathways. The drug-miRNA interaction analysis identified several repositioning opportunities. Metformin’s predicted interaction with miR-155 is particularly interesting given its established benefits in diabetic patients and could represent a direct mechanistic link.^21,22^

Laboratory-based validation protocols continue to represent the gold standard for therapeutic target confirmation, yet our integrated scoring methodology establishes a systematic framework for candidate prioritization in preclinical research pipelines. The multi-parametric assessment algorithm incorporates expression consistency, structural accessibility, network centrality, pathway significance, and druggability metrics through weighted scoring matrices. This computational triage approach optimizes resource allocation by stratifying miRNA targets according to translational potential before expensive experimental workflows. Bioinformatics-driven drug discovery increasingly relies on such composite evaluation systems to navigate the complexity of molecular target selection, particularly in polygenic diseases where traditional reductionist approaches prove inadequate for capturing systems-level therapeutic opportunities.

Several methodological constraints inherently limit the generalizability and precision of our study’s findings. Repository heterogeneity across genomic databases introduces systematic bias through varying experimental protocols, platform-specific technical artifacts, and inconsistent quality control standards that compromise meta-analytical robustness. Current target prediction algorithms exhibit substantial false-positive rates and limited sensitivity for non-canonical binding interactions, potentially overlooking clinically relevant miRNA-mRNA regulatory pairs. The static network reconstruction approach fails to capture temporal regulatory dynamics essential for understanding wound healing progression phases. Additionally, cross-species extrapolation assumptions may inadequately reflect human-specific miRNA processing mechanisms and target site evolution, necessitating species-matched experimental validation before therapeutic translation.

## CONCLUSION

This integrative bioinformatics analysis evaluated miRNA regulatory architecture in DFU pathophysiology, identifying eight consistently dysregulated miRNAs across multiple datasets and revealing modular network organization governing inflammation, angiogenesis, and extracellular matrix remodeling. The computational framework successfully prioritized therapeutic targets and identified promising drug repositioning candidates, establishing a systematic approach for miRNA-based therapeutic development in diabetic wound healing.

## Data Availability

All data produced in the present work are contained in the manuscript

